# Newborn Care Practices in Health Facilities of Nepal: A Further Analysis from Nepal Health Facility Survey 2015 and 2021

**DOI:** 10.1101/2023.05.26.23290601

**Authors:** Achyut Raj Pandey, Bikram Adhikari, Bipul Lamichhane, Bishnu Dulal, KC Saugat Pratap, Deepak Joshi, Sushil Chandra Baral

## Abstract

**Introduction:** Increased availability of Newborn care practices in health facilities (HFs) plays an important role in improving the survival and well-being of newborns. In this paper, we aimed to determine newborn care practices among HFs between 2015 and 2021, and associated factors among public and private HFs in Nepal.

**Methods:** We performed a secondary analysis of Nepal Health Facility Surveys 2015 and 2021. We summarized categorical variables with a weighted percent and 95% confidence interval (CI). We compared proportions using the z-test of proportion and reported differences in percentage points, 95% CI, and p-value. We applied univariate and multivariable logistic regression analysis to determine the association of the availability of seven newborn care practices.

**Results:** The percentage of facilities with all seven newborn care practices was 50.5% (95% CI: 44.6, 56.3) in 2015 and 83.7% (95%CI: 79.8, 87.0) in 2021 with an overall increase of 34.2 percent points (95% CI: 27.9, 38.7). The availability of all seven newborn care practices significantly increased from 2015 to 2021 in each subcategory of the ecological region, all provinces except Madhesh, and all types of HFs except federal/provincial hospitals. In 2021, private hospitals had lower odds of having all seven newborn care practices compared to federal/provincial hospitals (AOR= 0.17, 95% CI: 0.07, 0.04). Similarly, in 2021, Sudurpaschim province had 2.87 (95% CI: 1.06, 8.31) higher odds of having all seven newborn care practices compared to Koshi province. In 2021, newborn care practices did not differ significantly based on ecological belt, quality assurance activities, external supervision, delivery service-related training and frequency of HF meeting.

**Conclusion:** There was a significant increase in the availability of seven newborn care practices between 2015 and 2021 and in each category of ecological region, province, and type of facility. The type of facility and provinces were associated with the availability of newborn care practices among HFs in Nepal.

## Introduction

The first four weeks of life are still the most important for a child’s survival. Nearly 2.3 million children lost their lives in the first month of birth in 2021, which equates to about 6,400 fatalities every day [1]. Sub-Saharan Africa and Central and Southern Asia have the greatest mortality rates, while progress toward the worldwide 2030 objectives varies at different rates in all of these nations [2]. From 1990 to 2020, the worldwide Neonatal Mortality Rate (NMR) fell by more than 50%, from 37 to 17 deaths per 1,000 live births [1]. With a relatively slower pace of decline in NMR, the proportion of neonatal deaths out of total under 5 deaths increased from 39% in 2000 to 48% in 2019 [3, 4].

While countries around the globe are struggling to reduce the newborn mortality rate to 12 or fewer deaths per 1000 live births by 2030 as a part of Sustainable Development Goal (SDG) [5], the alarming and avoidable burden of newborn deaths raise concerns about SDGs’ success as over 60 countries are predicted to fall short of the goal for newborn mortality [1]. If NMR continues to decline in the current rate, there will be 13.9 newborn fatalities per 1,000 live births by 2030 [6], which is still more than the ideal NMR that can be achieved with the medical and technological advancements at hand. [6] According to an estimate based on predictions of the global burden of disease, the ideal worldwide NMR that could be achieved through medical and technological advancements is 0.80 deaths per 1,000 live births [3].

In concordance with global pattern, in Nepal, the decline in NMR has been relatively slow compared to the decline in under-five mortality rates. Between 2001 to 2022, under-5 mortality rates declined from 91 per 1000 live births to 33 per 100 live births in 2021, demonstrating a decline of approximately 64% in a period of two decades. [7, 8]. However, in the same period, NMR declined by 46% from 39 newborn deaths per 1000 live births in 2001 to 21 newborn deaths per 1000 live birth in 2022[7, 8]. By achieving the SDG, Nepal will avoid 5,935 impairments and save an additional 27,116 neonatal lives. Additionally, it has been demonstrated that programs aiming at improving maternal and newborn health are economically effective, with every dollar spent on newborn health yielding a return of $6 [9].

Newborns all over the world have the right to receive high-quality and essential newborn care which entails immediate medical attention after delivery and continued care throughout the newborn phase at the health facility level as well as at home [10]. It includes immediate care at birth, thermal care, resuscitation when needed, support for breast milk feeding, nurturing care, infection prevention, assessment of health problems, recognition and response to danger signs, and timely and safe referral when needed. Immediate care at birth involves delayed cord clamping, thorough drying, assessment of breathing, skin-to-skin contact, and early initiation of breastfeeding. Essential newborn care is necessary both in the health facility and at home to ensure the well-being and survival of newborns [10]. While the progress on newborn health was challenged by COVID-19 epidemic, rising poverty, and worsening humanitarian situations are adding to the strain on already overburdened healthcare institutions [2], health system in Nepal underwent a transformation from unitary to federal health system with substantial changes in health care delivery mechanism and the way health services are organized [11, 12]. In this context, we attempted to analyze the newborn care service practices, variation by province and ecological belts, change from 2015 to 2021, and factors associated with the availability of newborn care practices.

## Methods

### Study design

We used data from two nationally representative Nepal Health Facility Survey (NHFS) conducted in the year 2015 and 2021 for secondary data analysis. NHFS 2015 and NHFS 2021 were implemented by NEW Era, a national research firm with the support from Ministry of Health and Population (MOHP) and technical support from ICF International.

Nepal’s healthcare system operates at three levels: federal, provincial, and local. The Ministry of Health and Population (MoHP) at federal level oversees policy formulation, planning, coordination, and organization of healthcare at all levels. The MoHP regulates services and implements healthcare initiatives. The local health system comprises various facilities such as primary hospitals, PHCCs, health posts, basic health care centers, urban health clinics, community health units, and community-level health facilities, including community health clinics, primary health outreach clinics, and immunization clinics. Health posts or basic health service centers serve as the initial point of contact for basic health services. Each local level is supposed to have a local hospital as per policy provision after federalization process, which are currently being constructed. There are provincial and federal level services particularly secondary and tertiary level service in Nepalese context.

Each level beyond health posts serves as a referral point within a network that extends from PHCCs to primary and secondary level hospitals. Private health facilities, including hospitals, clinics, and pharmacies, deliver basic health services up to tertiary care.

### Sample and sampling techniques

In NHFS 2015, a total of 1000 HFs were selected out of 4719 eligible HFs using a random stratified sampling technique. A total of 963 health facilities (HFs) were included in the study after eliminating 8 duplicate HFs and accounting for 29 HFs that did not participate due to refusal, closure, or inaccessibility caused by poor infrastructure. NHFS 2021 utilized a stratified random sampling method to select 1633 health facilities from a pool of 5681 eligible facilities. Equal probability systematic sampling was employed, with sample allocation considering facility type within each province to achieve stratification. Out of the 1633 selected HFs, 97% (1576 facilities) successfully participated in the survey, while 2% were non-functional, 1% were unreachable, and 0.1% refused to take part in the study. In this study, we used data of 621 (weighted: 457) and 786 (weighted: 804) HFs offering normal vaginal delivery services from NHFS 2015 and NHFS 2021 respectively.

### Data collection

The NHFS 2015 data collection took place from April to November 2015, while the NHFS 2021 data collection occurred from January to September 2021. The data collection was carried out using four main data collection tools-a) facility inventory questionnaire, b) health provider interview questionnaire, c) observation for maternal and newborn care, and d) Exit interview questionnaire. We used data from the facility inventory questionnaire focusing on newborn care practices in this study.

### Variables

#### Outcome variable

Those facilities reporting presence of all seven newborn care practices – delivery to the abdomen, drying and wrapping, kangaroo mother care (KMC), initiation of breastfeeding within the first hour, routine complete head-to-toe examination of the newborn before discharge, applying chlorhexidine gel to umbilical cord stump and weighting the newborn immediately upon delivery were considered to have all newborn care practices.

### Independent variables

The independent variables included ecological region (hill/mountain/terai), location of facility (rural/urban), province (Koshi / Madhesh / Bagmati / Gandaki /Lumbini / Karnali / Sudurpaschim), type of facility (federal, provincial hospital / peripheral facilities / private hospital). The hospitals under federal or provincial government were classified as federal or provincial hospitals, facilities under local governments which include local hospitals, primary health care centers, and health posts were classified as local HFs, and the hospitals owned by private sectors were classified as private hospitals. For frequency of HFs meeting, the HFs stating “no” for routine management/administrative meetings were classified as “none”, those stating, “monthly or more often” were classified as “monthly” and those stating, “irregular or every 2-6 months” were classified as “sometimes”.

### Statistical analysis

We used R program 4.2.0 for data cleaning and statistical analysis. We performed a weighted analysis to address complex survey design using the “survey” package. In the descriptive analysis, we summarized categorical variables with weighted percent and 95% confidence interval. We compared proportions between 2015 and 2021 using the z-test of proportion and reported differences in percentage points, 95% CI, and p-value. In inferential analysis, we applied univariate and multivariable logistic regression analysis to determine the association of availability of seven newborn care practices with the province, type of HFs and ecological region. The results of regression analysis were reported with crude and adjusted odds ratio along with their 95% CI and p-value. We considered a p-value less than 0.05 as statistically significant throughout this paper.

### Ethical approval

The study involves a secondary analysis of NHFS 2015 and NHFS 2021 so, we did not seek ethical approval for this study. In the original survey NHFS 2015 and 2021, ethical clearance was obtained from the Ethical Review Board of Nepal Health Research Council and ICF International.

## Results

The distribution of HFs offering normal vaginal delivery services by ecological belt and province between NHFS 2015 and 2021 were similar. In 2021, 61.4% facilities were from hilly region, 17.0% facilities were from mountain region, and 21.7% facilities were from terai region whereas in NHFS 2015, 60.4% facilities were from hill, 14.8% were from mountain and 24.8% were from Terai in 2015. In 2021, 16.7% of facilities were from Koshi, 7.6% were from Madhesh, 18.8% were from Bagmati, 11.4% were from Gandaki, 16.9% were from Lumbini, 12.4% were from Karnali and 16.1% were from Sudurpaschim. Figure 1 also shows distribution of HFs in NHFS 2015 and NHFS 2021. The representation of HFs between the two surveys looks comparable.

**Figure 1:**
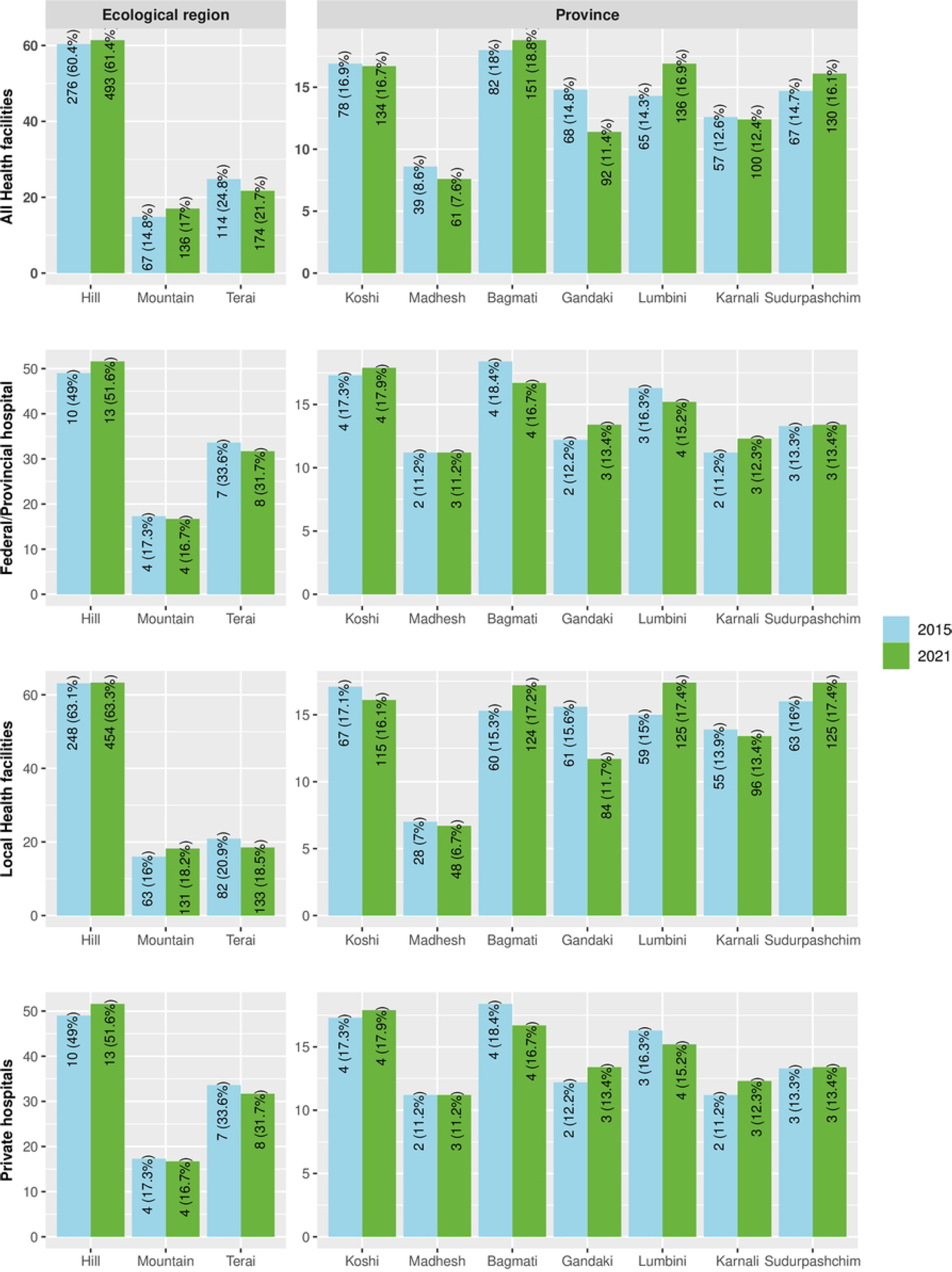
Distribution of facilities in NHFS 2015 and NHFS 2021

Table 1 presents the comparison of newborn care practices among HFs offering normal vaginal delivery services between NHFS 2015 and NHFS 2021. The availability of all seven newborn care practices was 49.5% (95% CI: 43.7, 55.4) in 2015 and 83.7% (95%CI: 79.8, 87.0) in 2021 with an overall increase of 34.2 percentage points (95%CI: 27.9, 38.7). In 2021, more than 90% facilities reported practicing delivery to the abdomen (96.1%, 95% CI: 93.7, 97.7), drying and wrapping of newborns (99.7%, 95% CI: 98.3, 99.9), early initiation of breastfeeding (99.4%, 95% CI: 98.1, 99.8), routine head-to-toe examination (97.4%, 95% CI: 95.3, 98.6), applying chlorhexidine gel to umbilical cord stump (96.5% 95% CI: 94.9, 97.6) and weighing the newborn immediately upon delivery (99.0%, 95% CI: 96.8, 99.7).

**Table 1:**
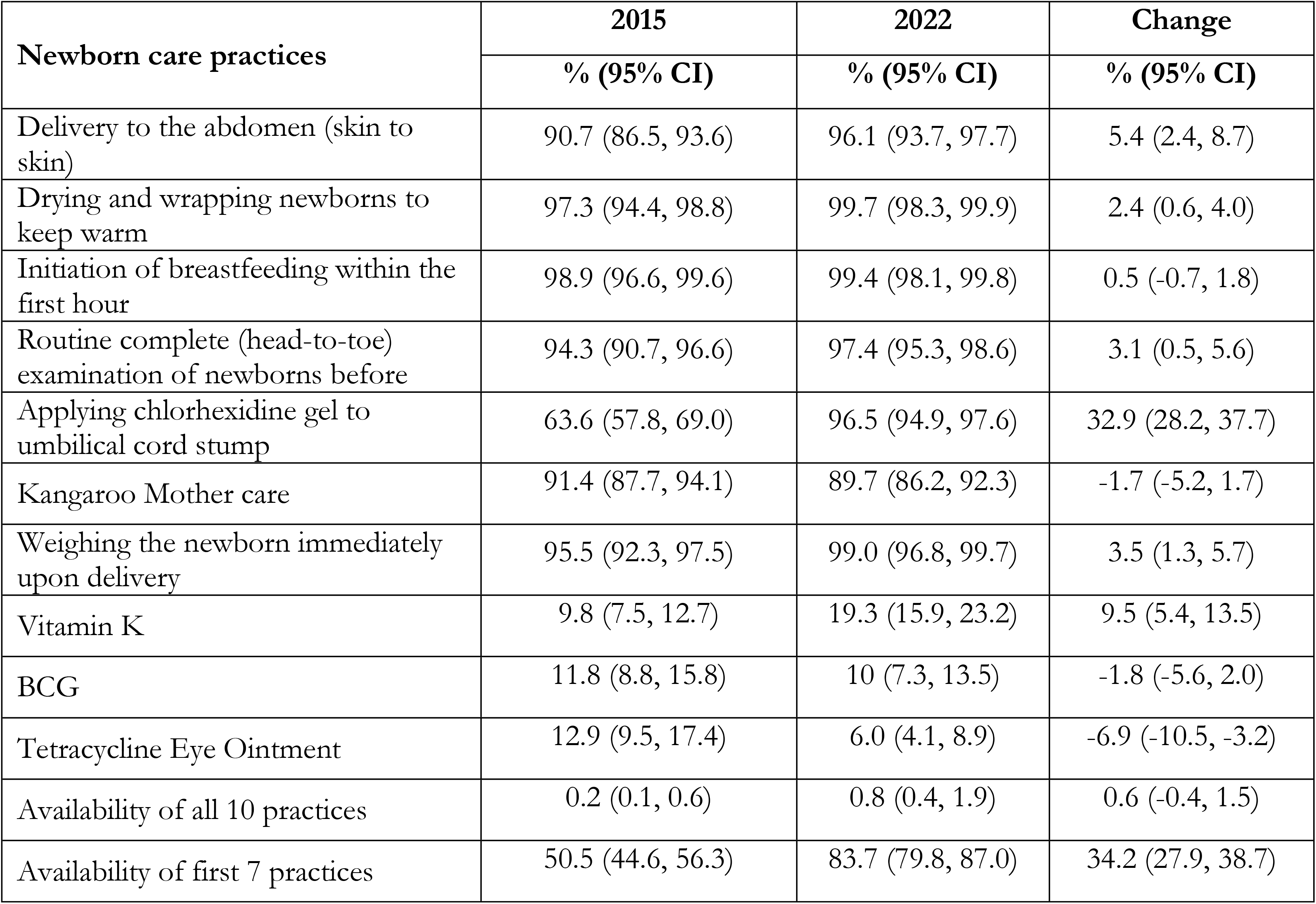
Comparison of availability of newborn care practices among HFs offering normal vaginal delivery between NHFS 2015 and NHFS 2021

The practice of applying chlorhexidine gel to umbilical cord stump increased from 63.6% (95% CI: 57.8, 69.0) in 2015 to 96.5% (95% CI: 94.9, 97.6) in 2021 with a total change of 32.4 percentage points (95% CI: 28.2, 37.7) and was the newborn care practice with highest percentage point improvement. The practice of drying or wrapping newborns, routine head-to-toe examination of newborns, weighing newborns immediately after delivery and Vitamin K use improved positively between 2015 to 2021. There were no significant changes in the practices of initiation of breastfeeding within the first hour, Kangaroo Mother Care (KMC), and BCG vaccination between 2015 and 2021.

The details of the comparison of availability of all seven newborn care practices NHFS 2015 and NHFS 2021 for different ecological belts and seven provinces stratified by type of HFs is present in the **supplementary table 1**.

Disaggregated by type of facilities, use of Vitamin K and application of chlorhexidine were the two indicators that improved most between 2015 and 2022 in federal and provincial level hospitals. Use of Vitamin K increased from 20.5% (95% CI: 13.5, 29.8) in 2015 to 65.1% (95% CI: 54.5, 74.4) in 2022 demonstrating 44.6 percentage points improvement. Similarly, in local level facilities, the use of chlorhexidine was the indicator that improved most increasing from 67.6% (95% CI: 61.0, 73.5) to 98.7% (95% CI: 96.8, 99.4) demonstrating 31.1 percentage points improvement. In private hospitals, the proportion of facilities applying chlorhexidine gel to umbilical cord stump increased from 25.4% (95% CI: 16.2, 37.4) to 72.3% (95% CI: 62.4, 80.4) with a total of 46.9 percentage points improvement. [Table 2]

**Table 2:** Newborn care practices by facility type

| Newborn care practices | Federal hospitals |  | Local facilities |  | Private hospitals |  |
| --- | --- | --- | --- | --- | --- | --- |
|  | 2015 | 2021 | 2015 | 2021 | 2015 | 2021 |
| Delivery to the abdomen (skin to skin) | 87.8<br>(79.5, 93.0) | 94.4<br>(87.0, 97.7) | 91.5<br>(86.8, 94.7) | 96.7<br>(94.0, 98.3) | 84.4<br>(71.3, 92.2) | 89.9<br>(79.4, 95.4) |
| Drying and wrapping newborns to keep warm | 100.0 | 98.9<br>(92.2, 99.8) | 97.2<br>(93.7, 98.8) | 100.0 | 97.0<br>(85.0, 99.5) | 95.9<br>(79.6, 99.3) |
| Initiation of breastfeeding within the first hour | 99.0<br>(92.9, 99.9) | 100.0 | 99.1<br>(96.1, 99.8) | 99.7<br>(98.5, 99.9) | 96.9<br>(85.0, 99.4) | 95.3<br>(80.7, 99.0) |
| Routine complete(head-to-toe) examination of newborns before | 99.0<br>(92.9, 99.9) | 95.5<br>(88.5, 98.3) | 94.2<br>(89.9, 96.7) | 97.9<br>(95.4, 99.0) | 94.0<br>(81.8, 98.2) | 93.0<br>(81.2, 97.6) |
| Applying chlorhexidine gel to umbilical cord stump | 70.4<br>(60.4, 78.7) | 93.3<br>(85.7, 97.0) | 67.6<br>(61.0, 73.5) | 98.7<br>(96.8, 99.4) | 25.4<br>(16.2, 37.4) | 72.3<br>(62.4, 80.4) |
| Kangaroo Mother care | 94.8<br>(88.0, 97.9) | 95.5<br>(88.4, 98.3) | 92.4<br>(88.1, 95.2) | 90.3<br>(86.5, 93.2) | 81.6<br>(69.1, 89.8) | 79.9<br>(70.0, 87.1) |
| Weighing the newborn immediately upon delivery | 100.0 | 100.0 | 95.3<br>(91.6, 97.5) | 99.2<br>(96.6, 99.8) | 95.3<br>(82.7, 98.8) | 96.4<br>(78.3, 99.5) |
| Vitamin K | 20.5<br>(13.5, 29.8) | 65.1<br>(54.5, 74.4) | 2.4<br>(1.2, 4.7) | 12.5<br>(9.2, 16.8) | 70.4<br>(57.0, 80.9) | 79.5<br>(69.6, 86.8) |
| BCG | 31.7<br>(23.1, 41.7) | 22.5<br>(14.9, 32.5) | 9.7<br>(6.5, 14.2) | 8.8<br>(6.0, 12.7) | 22.0<br>(13.2, 34.2) | 19.0<br>(11.1, 30.7) |
| Tetracycline Eye Ointment | 15.3<br>(9.4, 24.0) | 10.1<br>(5.3, 18.5) | 14.0<br>(10.1, 19.1) | 5.5<br>(3.3, 8.8) | 2.9<br>(1.2, 6.5) | 11.2<br>(7.1, 17.4) |
| Availability of all 10 practices | 3.1<br>(1.0, 9.2) | 2.2<br>(0.5, 8.7) | 0.1<br>(0.0, 0.4) | 0.7<br>(0.2, 2.0) | 0.5<br>(0.1, 3.6) | 1.7<br>(0.5, 5.3) |
| Availability of first 7 practices | 60.2<br>(50.0, 69.5) | 82.1<br>(72.5, 88.8) | 53.4<br>(46.7, 59.9) | 85.8<br>(81.4, 89.3) | 20.5<br>(12.4, 31.9) | 60.4<br>(50.2, 69.7) |

**Table 3:** Comparison of Availability of all seven newborn care practices between 2015 and 2021 by ecological region, province, and type of facility

| Characteristics | NHFS 2015 |  | NHFS 2015 |  | Change, 95% CI | P value |
| --- | --- | --- | --- | --- | --- | --- |
|  | n | % (95%CI) | n | % (95%CI) |  |  |
| **Ecological region** |  |  |  |  |  |  |
| Hill | 276 | 46.6(38.7, 54.7) | 493 | 86.1 (81.1, 89.9) | 39.5 (32.5, 46.4) | <0.001 |
| Mountain | 68 | 53.5(40.4, 66.2) | 136 | 83.2 (71.6, 90.7) | 29.7 (15.1, 44.2) | <0.001 |
| Terai | 114 | 58.0(47.8, 67.5) | 174 | 77.5 (68.0, 84.8) | 19.5 (7.8, 31.2) | 0.001 |
| **Province** |  |  |  |  |  |  |
| Koshi | 78 | 53.6 (39.9,66.7) | 134 | 76.6(64.4, 85.6) | 23 (8.8, 37.3) | 0.001 |
| Madhesh | 39 | 58.3(40.5,74.1) | 61 | 74.7 (55.1, 87.7) | 16.4 (-4.5, 37.4) | 0.131 |
| Bagmati | 82 | 36.9 (25.7, 50.0) | 151 | 89.7 (80.6, 94.8) | 52.8 (40.3, 65.2) | <0.001 |
| Gandaki | 68 | 50.2(32.5, 67.9) | 92 | 68.1 (52.5, 80.5) | 17.9 (1.3, 34.4) | 0.034 |
| Lumbini | 65 | 62.2(47.5, 74.9) | 136 | 89.3 (79.3, 94.8) | 27.1 (13.2, 41.1) | <0.001 |
| Karnali | 57 | 33.0(19.0,50.7) | 100 | 85.9 (74.9, 92.5) | 52.9 (37.6, 68.2) | <0.001 |
| Sudurpaschim | 67 | 62.6(48.2,75.1) | 130 | 91.9 (83.6, 96.2) | 29.3 (15.7, 42.9) | <0.001 |
| **Type of facility** |  |  |  |  |  |  |
| Federal/Provincial | 20 | 60.2(50.1,69.5) | 25 | 82.1(72.6, 88.8) | 21.9 (-8.8, 52.3) | 0.196 |
| Local HF's | 392 | 53.4(46.7,59.9) | 718 | 85.8(81.4, 89.3) | 32.4 (26.7, 38.2) | <0.001 |
| Private Hospitals | 45 | 20.5(12.5,31.8) | 61 | 60.4(50.2, 69.7) | 9.9 (20.9, 58.8) | <0.001 |

**Table 4:** Association of factors with availability of all newborn care practices except BCG, tetracycline, and Vitamin-K

| Characteristic | 2015 |  |  |  | 2021 |  |  |  |
| --- | --- | --- | --- | --- | --- | --- | --- | --- |
|  | Unadjusted |  | Adjusted |  | Unadjusted |  | Adjusted |  |
|  | OR (95%CI) | p-value | OR (95% CI) | p-value | OR (95%CI) | p-value | OR (95% CI) | p-value |
| **Ecological Region** |  |  |  |  |  |  |  |  |
| Hill | Ref |  | Ref |  | Ref |  | Ref |  |
| Mountain | 1.32 (0.71, 2.44) | 0.378 | 1.45 (0.75, 2.79) | 0.266 | 0.8 (0.37, 1.72) | 0.572 | 0.64 (0.26, 1.54) | 0.316 |
| Terai | 1.58 (0.94, 2.66) | 0.086 | 1.55 (0.78, 3.07) | 0.213 | 0.56 (0.30, 1.02) | 0.058 | 0.59 (0.26, 1.34) | 0.210 |
| **Province** |  |  |  |  |  |  |  |  |
| Koshi | Ref |  | Ref |  | Ref |  | Ref |  |
| Madhesh | 1.21 (0.49, 2.96) | 0.678 | 0.96 (0.35, 2.68) | 0.945 | 0.9 (0.32, 2.57) | 0.843 | 1.2 (0.37, 3.92) | 0.763 |
| Bagmati | 0.51 (0.24, 1.09) | 0.081 | 0.56 (0.24, 1.31) | 0.183 | **2.65 (1.03, 6.79)** | **0.042** | 2.53 (0.93, 6.90) | 0.069 |
| Gandaki | 0.87 (0.35, 2.17) | 0.771 | 0.88 (0.33, 2.35) | 0.804 | 0.65 (0.27, 1.57) | 0.336 | 0.46 (0.18, 1.22) | 0.119 |
| Lumbini | 1.42 (0.64, 3.18) | 0.391 | 1.41 (0.59, 3.39) | 0.441 | **2.54 (0.96, 6.73)** | **0.060** | 2.22 (0.80, 6.14) | 0.125 |
| Karnali | 0.43 (0.17, 1.06) | 0.065 | 0.41 (0.16, 1.03) | 0.058 | 1.85 (0.74, 4.64) | 0.187 | 1.42 (0.52, 3.92) | 0.497 |
| Sudurpaschim | 1.45 (0.65, 3.22) | 0.362 | 1.26 (0.55, 2.87) | 0.586 | **3.45 (1.28, 9.29)** | **0.014** | **2.87 (1.03, 7.99)** | **0.044** |
| **Type of Facility** |  |  |  |  |  |  |  |  |
| Federal/Provincial hospital | Ref |  | Ref |  | Ref |  | Ref |  |
| Local level facilities | 0.76 (0.47, 1.23) | 0.263 | 0.81 (0.47, 1.42) | 0.469 | 1.32 (0.70, 2.48) | 0.393 | 1.09 (0.53, 2.26) | 0.811 |
| Private Hospital | **0.17 (0.08, 0.35)** | **<0.001** | **0.17 (0.07, 0.40)** | **<0.001** | **0.33 (0.17, 0.66)** | **0.002** | **0.26 (0.11, 0.63)** | **0.003** |
| **Quality assurance activities** |  |  |  |  |  |  |  |  |
| Not Performed | Ref |  | Ref |  | Ref |  | Ref |  |
| Performed | 0.7 (0.42, 1.19) | 0.189 | 0.57 (0.31, 1.03) | 0.062 | 1.1 (0.61, 1.98) | 0.75 | 1.01 (0.52, 1.94) | 0.983 |
| **External supervision** |  |  |  |  |  |  |  |  |
| No | Ref |  | Ref |  | Ref |  | Ref |  |
| Yes | 1.41 (0.83, 2.41) | 0.207 | 1.15 (0.65, 2.05) | 0.626 | 1.55 (0.90, 2.68) | 0.112 | 1.42 (0.78, 2.57) | 0.248 |
| **Provider with training on delivery** |  |  |  |  |  |  |  |  |
| No staff with training | Ref |  | Ref |  | Ref |  | Ref |  |
| At least one staff with training | **1.78 (1.08, 2.91)** | **0.023** | 1.4 (0.83, 2.36) | 0.207 | 0.88 (0.48, 1.60) | 0.666 | 0.69 (0.35, 1.36) | 0.285 |
| **Health facility meeting frequency** |  |  |  |  |  |  |  |  |
|  | Unadjusted |  | Adjusted |  | Unadjusted |  | Adjusted |  |
|  | OR (95%CI) | p-value | OR (95% CI) | p-value | OR (95%CI) | p-value | OR (95% CI) | p-value |
| None | Ref |  | Ref |  | Ref |  | Ref |  |
| Sometimes | **2.28 (1.00, 5.18)** | **0.049** | **2.45 (1.04, 5.76)** | **0.040** | 1.53 (0.59, 3.98) | 0.379 | 1.39 (0.50, 3.87) | 0.533 |
| Monthly | **2.21 (1.19, 4.09)** | **0.012** | **2.1 (1.09, 4.04)** | **0.026** | 1.44 (0.65, 3.19) | 0.371 | 1.36 (0.58, 3.21) | 0.477 |
| OR = Odds Ratio, CI = Confidence Interval |  |  |  |  |  |  |  |  |

Figure 2 shows the proportion of facilities reporting seven newborn care practices in terms of scores from one to seven. A score of one means that the facility has one out of seven newborn care practices while seven indicates that the facility has all seven newborn care practices. In 2021, 83.7% of HFs reported having all seven newborn care practices which is an increase from 50.5% in 2015. Disaggregated by facility type, the proportion of facilities reporting all seven newborn care practices increased from 60.2% in 2015 to 82.1% in 2021 in federal or provincial level hospitals, 53.4% in 2015 to 85.8% in 2022 in local HFs and, from 20.5% in 2015 to 60.4% in 2022 in private hospitals.

**Figure 2:**
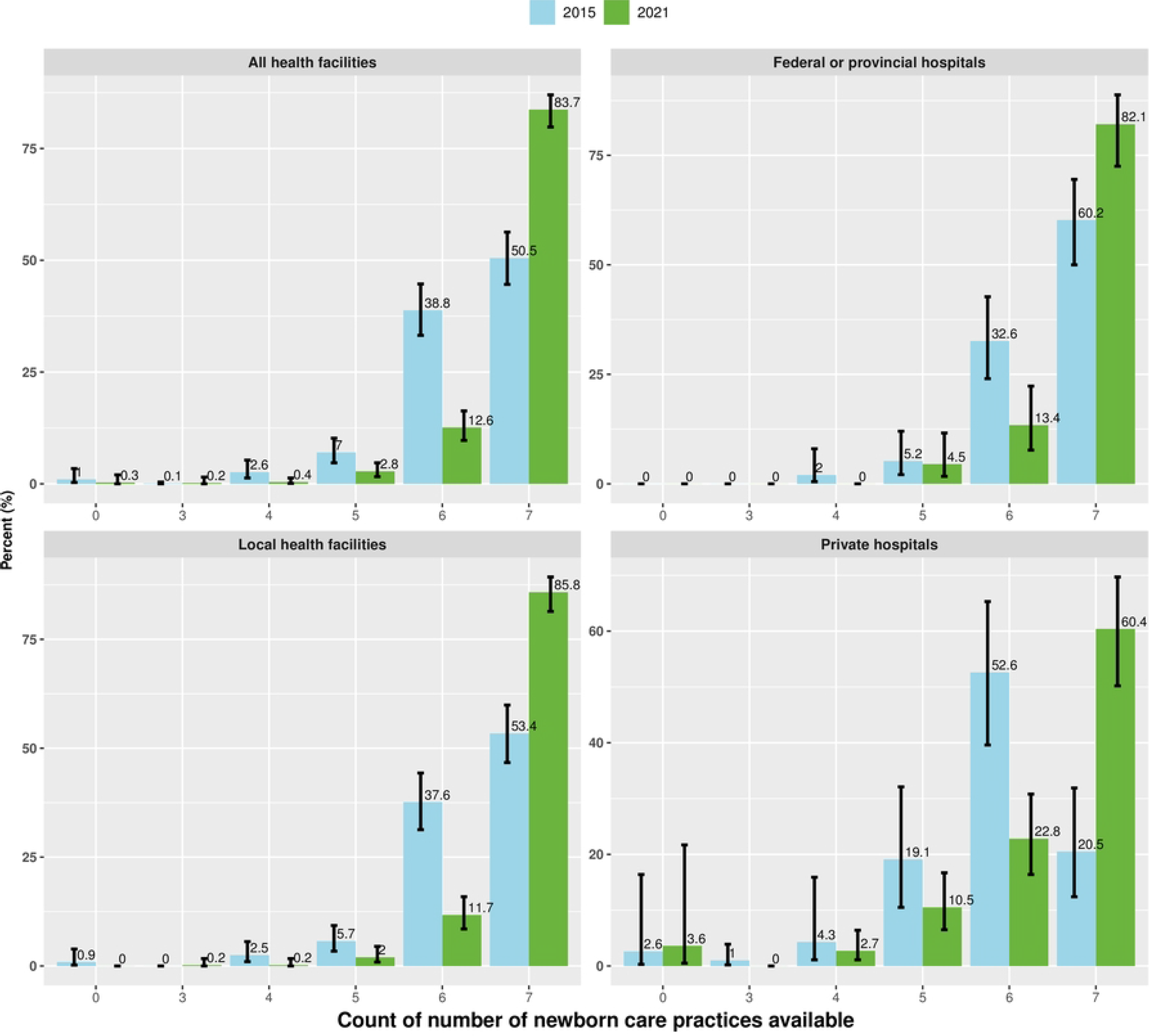
Comparison of availability newborn care practices score between NHFS 2015 and NHFS 2021

Among ecological belts, the hill region witnessed the highest percentage point improvement in the practice of all seven newborn signal functions. In the hilly region, the proportion of facilities that reported practicing all seven newborn care practices increased from 46.6 % (95% CI: 38.7, 54.7) in 2015 to 46.6 % (95% CI: 38.7, 54.7) in 2021 demonstrating a change of 39.5 percentage points (95% CI: 32.5, 46.4). Among provinces, improvement was seen in Karnali and Bagmati Province. In Karnali province, percentage of facilities with all newborn care practices increased from 33.0% (95% CI: 19.0,50.7) in 2015 to 85.9% (95% CI: 74.9, 92.5) in 2021 demonstrating a change of 52.9 percentage points (95% CI: 37.6, 68.2). Similarly, In Bagmati province, the percentage of facilities with all seven newborn care practices improved from 36.9% (95% CI: 25.7, 50.0) in 2015 to 89.7% (95% CI: 80.6, 94.8) in 2021 demonstrating a change of 52.8 percentage points (95% CI: 40.3, 65.2)

In multivariate analysis in Table 5, the odds of availability of all newborn care practices in the year 2015 was 0.17 (95% CI: 0.07, 0.04; p-value=<0.001) times in private hospitals compared with federal/provincial hospitals, 2.45 (95% CI: 1.04, 5.76, p-value: 0.040) times in HFs having health facility meeting for sometimes and 2.1 (95% CI: 1.03, 7.99, p-value:0.044) times in facilities having monthly health facility meeting compared to HFs with no health facility meeting. The odds of having all seven newborn care practices were 2.87 (95% CI: 1.06, 8.31; p-value: 0.038) times in Sudurpaschim province compared to Koshi. Similarly, the odds of having all seven newborn care practices were 0.26 (0.11, 0.63; p-value=0.003) times in private facilities compared to federal/provincial hospitals

## Discussion

The availability of all newborn care practices has increased except kangaroo mother care between 2015 and 2021. The availability of seven newborn care practices increased to 84% in 2021 from 50% in 2021 with an overall increase of 34% point. In 2021 and 2015, private hospitals had lower odds of having all seven newborn care practices compared to federal/provincial hospitals. In 2021, Sudurpaschim province had 3 times higher odds of having all seven newborn care practices compared to Koshi province. Availability of seven newborn care practices did not differ significantly based on ecological belt, quality assurance activities, external supervision, delivery service-related training and frequency of HF meeting.

Furthermore, HFs that held meetings on a monthly or occasional basis were found to have higher odds of having all seven practices. This finding underscores the need for regular collaboration and communication among health facility staff to promote the availability of essential newborn care practices.

In the regression model, with reference to Koshi province, Sudurpaschim province had almost 3-fold higher odds of practicing all seven essential newborn care practices. Between 2015 to 2019, the proportion of facilities practicing all seven essential newborn care practices increased by 29%. Notable positive changes in coverage of essential newborn care practices in Sudurpaschim province could be because of the higher priority placed in the health sector compared to other provinces. For example, the per capita spending on health increased from 607 per capita to 2941 per capita in Sudurpaschim province demonstrating approximately 3.8 folds increase while it increased from 1821 to 3432 at the national level which reflects around 88% increase in the same period [7].

The proportion of facilities performing all 7 signal functions for newborns increased from 50% to 84% with almost 34 percentage points increase. This progress has been driven by an increase in coverage of increase in the proportion of HFs applying chlorhexidine gel to umbilical cord stump which increased from 64% to 97%, with almost 33 percentage points increase in the period. Based on the assessment conducted in 10 districts, the government of Nepal decided to scale up the chlorhexidine programme to 41 districts by 2014 [13]. In the first phase of chlorhexidine programme between October 2011 and September 2014, 49 applications of chlorhexidine was implemented in 49 districts which were then gradually scaled up throughout the country after October 2014.[14] By Mid of 2016, 58 districts were implementing chlorhexidine at health facility and community levels by mid-2016 which has not been scaled up throughout the country [13, 15]. The jump in the proportion of HFs applying chlorhexidine gel to the umbilical cord stump could be because of policy changes or the decision of the government of Nepal to scale up the programme throughout the country.

Despite the high importance placed on the KMC, the proportion of facilities practicing KMC declined from 61% to 86% between 2015 to 2021. The KMC approach encompasses several interventions, such as continuous and early skin-to-skin contact, assistance with breastfeeding, early hospital discharge, and supportive care for stable neonates. Implementing this care package in hospitals for preterm neonates weighing less than 2000 g has been identified to decrease the risk of neonatal mortality by 51% [16], hypothermia by 77% [17], and also can shorten the duration of hospital stay. Although 86% of facilities reported adopting KMC, the coverage seems to decline further when it is captured at population level as all newborns delivered or visiting facilities with KMC may not receive the service. Multiple Indicators Cluster Survey in 2019 reported that the proportion of babies receiving KMC service was a cost-effective intervention to care for stable preterm/low birth weight (LBW) babies that is being implemented by the Government of Nepal as special care for small and/or sick new-born. Multiple Indicators Cluster Survey in 2019 reported that the proportion of babies receiving KMC service was a cost-effective intervention to care for stable preterm/low birth weight (LBW) babies that is being implemented by the Government of Nepal as special care for small and/or sick new-born. Although skin –to-skin contact has been part of different programs/ training packages such as Community-Based Integrated Management of Neonatal and Childhood Illnesses (CB-IMNCI), Facility-Based Integrated Management of Neonatal and Childhood Illnesses (FB-IMNCI), Skilled Birth Attendants (SBA) training and Comprehensive level-II new-born care, a full-fledged KMC program was rolled out in the year 2021 [13].

Despite 30 percentage point increment in proportion of facilities with all seven signal functions available, the latest evidence from Nepal Demographic and Health Survey (NDHS) 2022 show that NMR has remained stagnant at 21 per 100,000 live births between 2016 to 2021[7, 15]. As discussed above, this might be because chlorhexidine has substantial contribution in driving the aggregate figures upward which has limited contribution in reducing the NMR in Nepalese context. There are evidence with moderate certainty that using chlorhexidine on the umbilical cord stump as a routine practice does not appear to significantly impact neonatal mortality when compared to dry cord care or usual care [18]. WHO has set context specific recommendations regarding use of chlorhexidine, recommending only in setting where harmful traditional substances (e.g. animal dung) are commonly used on the umbilical cord [19].

Our study suggests that a relatively lower proportion of private hospitals tend to practice all seven signal functions for newborns compared to federal/provincial hospitals. Private health facility has important role in maternal and newborn health service delivery in Nepal. For example, as per NDHS 2016, out of total 57.3% deliveries take place in HFs; 10.2% deliveries take place in private facilities. Similarly, out of 56.8% newborns who received first postnatal checkup within 2 days of birth, 40.6% had received such service from government while the contribution of private sector was 9.8% which indicate the need of involving private sector in expanding quality newborn service coverage and improving neonatal health outcomes [15]. However, the lower proportion of private hospitals practicing all seven newborn care practices has raised concerns on compliance to standard protocol and quality of service. The Government should set strategies to regularly monitor and supervise private HFs relating to newborn service delivery. Steps may be taken for capacity development of private HFs through organizational twining, whereby, public hospitals with better compliance to newborn care practices and yielding better health outcomes may coach or engage selected private hospital within the province to improve compliance and quality of care. Regular interaction with private sector service providers, periodic discussion on policy provisions and facilitating them to develop infrastructures required for newborn services could be useful.

There are several strengths of this study. First, data collection for NHFS 2015 and NHFS 2021 was carried out in nationally representative samples, so the finding of this study is generalizable to Nepalese context. Second, NHFS 2015 and 2021 used validated tools and availability of tracer items or services are based on the observation by the enumerator thus ensuring the internal validity. Third, it is the first study based on nationally representative sample of HFs that allows us to compare the progress in newborn care practices after Nepalese health care delivery system underwent federalization process.

Despite several strengths, there are two major limitations to this study. First, we utilized data from a NHFS 2021 that was designed to assess the service availability and readiness for multiple services like antennal care, family planning, family planning, non-communicable diseases related services and so on and was not primarily dedicated to essential newborn care so some important variables like number of deliveries, newborn complications that could provide better context to the findings are missing. Second, the data collection of NHFS 2021 was undertaken during the period of COVID-19 pandemic, newborn care practices may have been under-estimated or over-estimated in NHFS 2021 owing to changes in service organization during the pandemic period.

## Conclusion

The availability of seven newborn care practices has increased between 2015 and 2021 and was relatively lower in private hospitals compared to federal or provincial hospitals. Further scale up and quality improvement of newborn care practices by HFs is essential to improve newborn survival and well-being.

## Data Availability

The data is available publicly in the open-access repository. The data can be downloaded from the official website of “The Demographic and Health Surveys” program. NHFS2021: https://dhsprogram.com/data/dataset/Nepal_SPA_2021.cfm?flag=0 NHFS2015: https://dhsprogram.com/data/dataset/Nepal_SPA_2015.cfm?flag=1)

## Acknowledgement

We would like to acknowledge DHS program for providing us data for further analysis and we are grateful to those who directly or indirectly contributed to this study and motivated us to conduct this study.

## Patient consent for publication

Not applicable, the article involves secondary analysis of survey data.

## Data availability statement

The data is available publicly in the open-access repository. The data can be downloaded from the official website of “The Demographic and Health Surveys” program.

*NHFS2021:* https://dhsprogram.com/data/dataset/Nepal_SPA_2021.cfm?flag=0

*NHFS2015:* https://dhsprogram.com/data/dataset/Nepal_SPA_2015.cfm?flag=1)

